# Stepping out of lockdown should start with school re-openings while maintaining distancing measures. Insights from mixing matrices and mathematical models

**DOI:** 10.1101/2020.05.12.20099036

**Authors:** Emma S McBryde, James M Trauer, Adeshina Adekunle, Romain Ragonnet, Michael T. Meehan

**Affiliations:** Australian Institute of Tropical Health and Medicine, James Cook University; Monash University

## Abstract

Australia is one of a few countries which has managed to control COVID-19 epidemic before a major epidemic took place. Currently with just under 7000 cases and 100 deaths, Australia is seeing less than 20 new cases per day. This is a positive outcome but makes estimation of current effective reproduction numbers difficult to estimate. Australia, like much of the world is poised to step out of lockdown and looking at which measures to relax first.

We use age-based contact matrices, calibrated to Chinese data on reproduction numbers and difference in infectiousness and susceptibility of children to generate next generation matrices (NGMs) for Australia. These matrices have a spectral radius of 2.49, which is hence our estimated basic reproduction number for Australia. The effective reproduction number *(R_eff_)* for Australia during the April/May lockdown period is estimated by other means to be around 0.8. We simulate the impact of lockdown on the NGM by first applying observations through Google Mobility Report for Australia at 3 locations: home (increased contacts by 18%), work (reduced contacts by 34%) and other (reduced contacts by 40%), and we reduce schools to 3% reflecting attendance rates during lockdown. Applying macro-distancing to the NGM leads to a spectral radius of 1.76. We estimate that the further reduction of the reproduction number to current levels of *R_eff_* = 0.8 is achieved by a micro-distancing factor of 0.26. That is, in a given location, people are 26% as likely as usual to have an effective contact with another person.

We apply both *macro* and *micro-distancing* to the NGMs to examine the impact of different exit strategies. We find that reopening schools is estimated to reduce *R_eff_* from 0.8 to 0.78. This is because increase in school contact is offset by decrease in home contact. The NGMs all estimate that adults aged 30-50 are responsible for the majority of transmission. We also find that *micro-distancing* is critically important to maintain *R_eff_* <1. There is considerable uncertainty in these estimates and a sensitivity and uncertainty analysis is presented.

## Introduction

Australia is now sustaining an estimated effective reproduction number (R_eff_) less than one across all jurisdictions. This success has prompted government to consider easing current lockdown restrictions, with several industries set to be re-opened. Among these are schools – whose shutdown has been an unceasing source of controversy.

Despite consistent advice to Commonwealth that childhood disease poses little risk for community transmission, most jurisdictions have enacted school closure policies. In view of the strategy of easing current restrictions whilst maintaining a suppression trajectory ^1^, this paper aims to address in epidemiological and modelling terms, the impact of opening schools, returning to workplaces and easing home lockdown requirements on the control of the COVID-19 epidemic in Australia.

### Methods

To estimate the relative contributions to transmission provided by different locations we begin with national age-specific contact rates generated by Prem *et al*. ^2^ (Figure 1). Initially, we weight these contact rates according to the relative susceptibility and infectiousness of different age groups, before multiplying through by the mean transmission rate per contact over the lifetime of infection. Matrix scaling factors are calibrated to the age-distribution of cases and basic reproduction number *R_0_* = 2.68 for China at baseline ^3,4^ We then applied the calibrated values to the Australian contact matrix, and estimated R_0_ for Australia.

**Figure 1.**
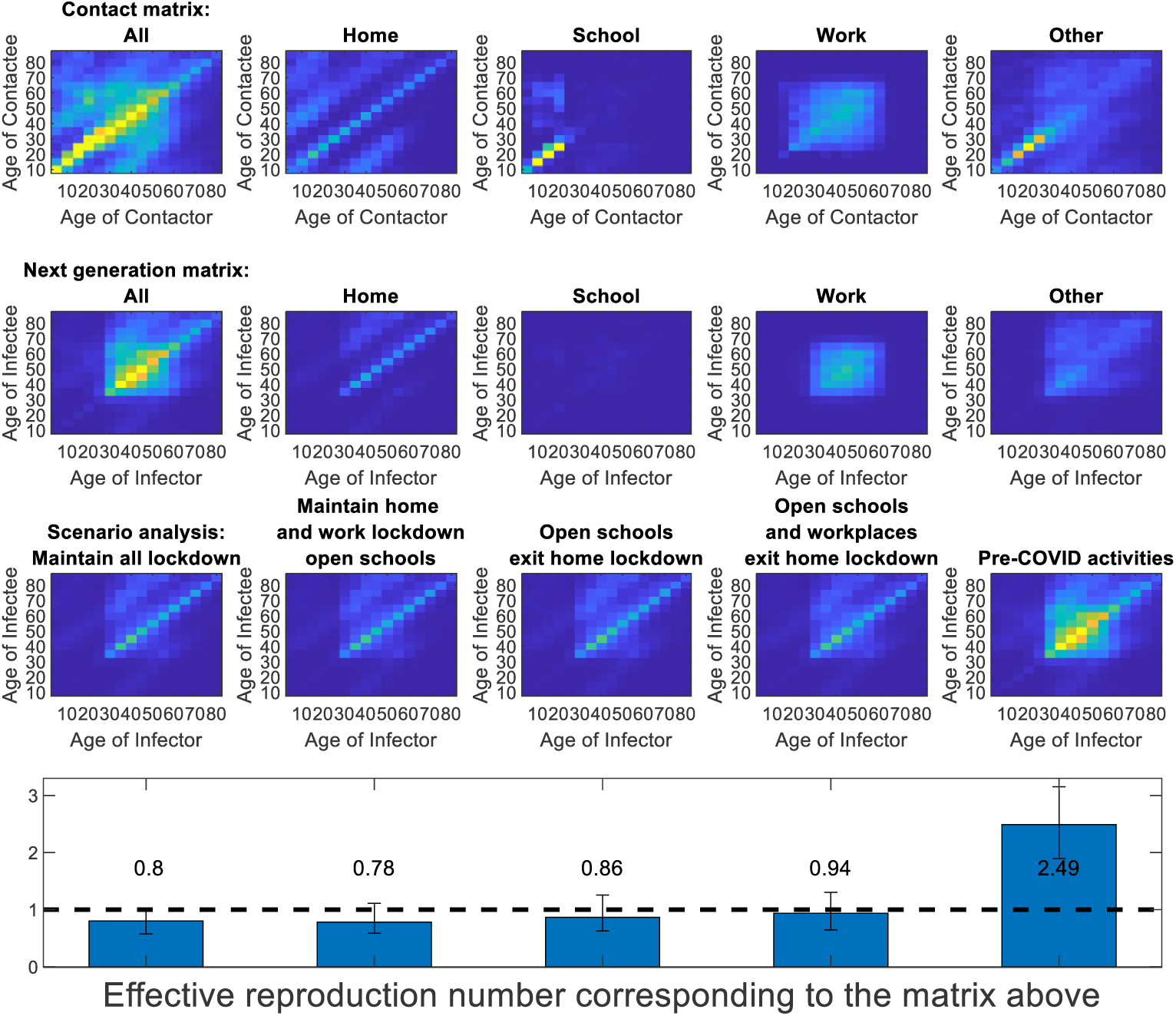
Top row of panels, mixing matrices by 5-year age groups for Australia taken from Prem *et al*.,^2^ overall and by four “location” components. Second row of panels, next generation matrices for each location (see SOM for derivation). Third row of panels, impact of five scenarios on the next generation matrix. Fourth row of panels, resulting *R_eff_*. All four intervention scenarios assume that micro-distancing is sustained at current high levels. The final value shows the impact of removing micro-distancing and returning to contact rates pre-COVID-19.

We apply Commonwealth and Google data to adjust location-specific mixing subsequent to restrictive policies and further scale for behavioural *“micro-distancing”* to reach Australia’s lockdown estimated *R_eff_* of 0.8. ^5^

We examined five intervention scenarios and the impact on *R_eff_*.

1. Current conditions continued
2. Return to school
3. Return to school and cease home lockdown, but with *micro-distancing*
4. Return to school and work, and cease home lockdown, but with improved micro-distancing
5. Return to pre-COVID-19 activity levels

See supporting online materials (SOM) for elaboration.

### Results

We estimate *R_0_* for Australia to be 2.49. Figure 1 demonstrates the contact and infection matrices under five scenarios. We estimate that schools opening while maintaining the current lockdown reduces *R_eff_* from 0.8 to 0.78, exiting home lockdown leads to *R_eff_* = 0.86, and work returning leads to *R_eff_* = 0.94, provided strenuous levels of micro-distancing remain in place. Outcomes are sensitive to model parameters, explored in the SOM.

### Discussion

We use Australian age-specific contacts, and global COVID-19 infection rates to estimate the impact of government policies on Australian COVID-19 control. We estimate a slightly lower *R_0_* for Australia than for China due to differences in mixing patterns. We then calibrate the impact of interventions undertaken through March-May 2020 to an effective reproduction number of 0.8. Our contact data-derived matrices show that age-assortative mixing between adults at home, work and the community are the major sources of transmission.

We examine strategies to move out of lockdown and conclude that school-opening would have minimal impact on COVID-19 transmission and is compensated by reduced home transmission. Changes in out-of-home contacts have greater impact. Although the role of micro-distancing remains highly uncertain, reopening workplaces and ceasing lockdown while sustaining strict distancing may allow suppression of COVID-19.

## Data Availability

data used are publically available

code is provided

